# Relationship between differences in physical activity patterns and blood glucose variability under free-living conditions in adults without diabetes

**DOI:** 10.64898/2026.11.25.26368827

**Authors:** Hyeon-Ki Kim, Yuka Hamada, Yoichi Hatamoto, Mana Hatanaka, Hinako Nanri, Takashi Nakagata, Yi-An Chen, Naoyuki Matsumoto, Seiya Shimoda, Shigeho Tanaka, Motohiko Miyachi, Yu Tahara, Shigenobu Shibata, Eiichi Yoshimura

## Abstract

The purpose of this study was to examine the relationship between differences in physical activity patterns and blood glucose variability under free-living conditions in healthy adults. A total of 102 adult participants (60 women, 42 men; age ≥20 years) wore a triaxial accelerometer for one week to record physical activity. Participants were then categorized into three groups based on the predominant time of day of physical activity: morning (06:00-12:00), afternoon (12:00-18:00), and night (18:00-24:00). Continuous glucose monitoring was used to assess blood glucose variability. Additionally, a prescribed breakfast was served on Day 1 of the experiment, and blood glucose fluctuations were assessed up to 2 hours after consumption. 24-hour blood glucose variability differed according to physical activity patterns, with the evening group having a lower 24-hour blood glucose variability than the morning and afternoon groups. In addition, the night group (127.7 ± 9.4 ml/dl) had significantly lower values 45 minutes after the prescribed meal than the morning (149.3 ± 9.5 ml/dl) and afternoon groups (146.7 ± 5.6 ml/dl) (p < 0.05). These findings suggest that engaging in physical activity during the evening is associated with reduced daily blood glucose fluctuations and smaller post-meal glucose spikes in healthy adults.

---

The prevalence of type 2 diabetes has dramatically increased over the past few decades ^1^. According to the International Diabetes Federation Report 2021, approximately 537 million adults (20–79 years) around the world have diabetes ^1^. Preventing and improving diabetes is important because the number of people with diabetes may become increasingly problematic in the future.

Physical activity is a key component in managing and controlling blood glucose levels, especially for individuals at risk of developing type 2 diabetes ^2^. Regular physical activity has been reported to be associated with a lower incidence of diseases, such as type 2 diabetes and cardiovascular disease ^3,4^. In humans, glucose tolerance shows diurnal variations, and has been reported to be lower in the evening compared to the morning ^5^. Insulin sensitivity and β-cell responsiveness to glucose, which are thought to contribute to the decline in glucose tolerance, were also lower in the evening compared to the morning ^6^. Therefore, blood glucose levels tend to be higher in the evening than in the morning. Thus, the effects of physical activity patterns on blood glucose levels may differ. Previous studies have shown that metabolic responses to exercise vary with the time of the day ^7–9^. Several intervention studies have shown that the timing of exercise regulates weight loss ^10,11^, blood glucose levels, and blood lipids ^12,13^. However, these studies evaluated the effects of constructed exercise timings. In contrast, much less is known about the influence physical activity patterns on glycemic variability under free-living conditions. In daily life, physical activity may take the form of housework and commuting. Therefore, it is important to investigate the relationship between the temporal distribution of physical activity accumulated throughout the day, and its health benefits in humans.

To date, most studies on the relationship between physical activity patterns and blood glucose levels have only focused on moderate-to-vigorous physical activity (MVPA) ^14–17^. Accumulating more MVPA in the evening between daily activities has been reported to have beneficial effects on glucose homeostasis in sedentary adults with overweight/obesity and metabolic disorders ^14^. Moreover, increased physical activity has been reported to be beneficial for glycemic control ^18,19^. Therefore, it is necessary to consider not only MVPA but also the number of steps in physical activity. Furthermore, humans spend most of the day in the postprandial state ^20^, whereas previous studies have primarily focused on fasting blood glucose levels ^15^. In addition, the balance of nutrient intake has been shown to be associated with the risk of type 2 diabetes ^21^. However, dietary intake has not been considered in previous studies examining the association between physical activity patterns and glycemic variability ^14–17^. Therefore, an evaluation that includes dietary controls is needed. Moreover, using continuous glucose monitoring to assess 24-hour blood glucose fluctuations can provide more detailed insights and is essential for evaluating the relationship between physical activity patterns and glycemic control ^22–24^.

Therefore, the purpose of this study was to examine the relationship between differences in physical activity patterns (number of steps and MVPA) and blood glucose variability under free-living conditions in healthy adults. Additionally, to account for the effects of diet on blood glucose levels, it used a prescribed diet.

## Results

Fig. 1 shows the experimental design of this study.

**Figure 1.**
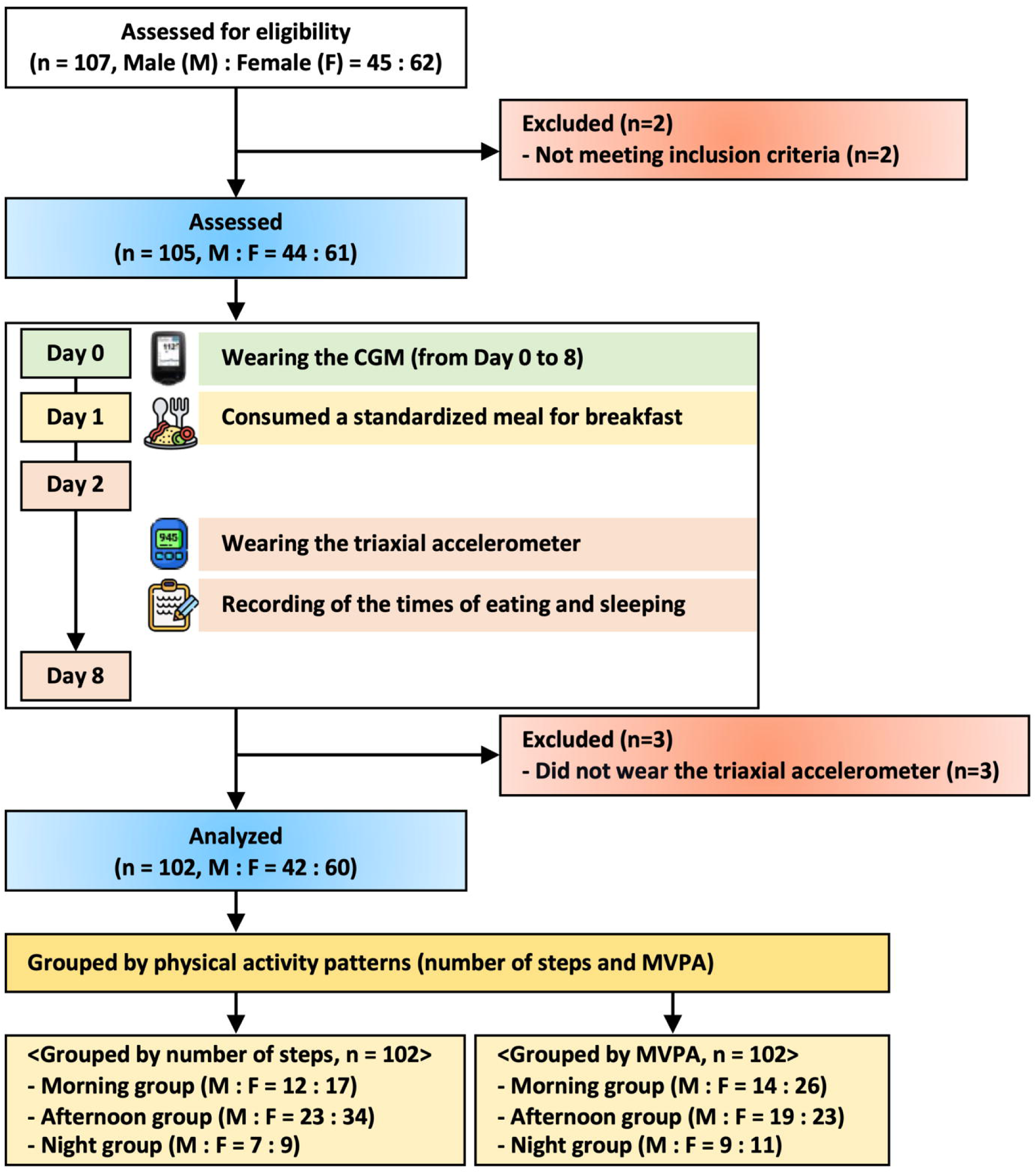
Flow diagram of the study participants. CGM: continuous glucose monitoring, MVPA: moderate-to-vigorous physical activity.

### Characteristics of participants

The characteristics of the 102 participants are shown in Table S1. Males had significantly higher height, weight, and BMI than females (p < 0.001, p < 0.001, and p = 0.023, respectively). Dietary intake also differed between females and males, with total energy, protein, fat, carbohydrate (p < 0.001), and dietary fiber intake (p = 0.009) being significantly higher in males than in females. However, no statistically significant differences were observed between males and females for physical activity, sleep, and meal times.

To examine the relationship between physical activity patterns and blood glucose variability, participants were divided into groups according to step count and MVPA. The characteristics of each group are shown in Tables S2 to S7. The physical activity pattern by step count showed that the morning group comprised significantly older participants than the afternoon and night groups (Table S2; p < 0.05, both). In terms of physical activity levels, the morning group had a significantly higher proportion of morning steps than the afternoon and night groups (Table S2; p < 0.001, both), and a higher proportion of morning MVPA (Table S2; p < 0.001, both). The afternoon group had significantly higher proportions of afternoon steps and MVPA than the morning and night groups (p < 0.001, both). The night group had significantly higher proportions of night steps and MVPA (Table S2; p < 0.001, both) as well as a significantly later bedtime than the morning and afternoon groups (Table S2; p < 0.001, both). However, no statistically significant difference in dietary intake was found among the groups. When the characteristics of each group were examined separately for males and females, trends similar to those in the overall results were found (Tables S3 and S4).

Next, when examining physical activity patterns using MVPA, those in the morning group were found to be significantly older than those in the afternoon and night groups (Table S5; p < 0.01, both). Regarding physical activity levels, the morning group had a significantly higher proportion of morning steps and MVPA than the afternoon and night groups (Table S5; p < 0.001, both). The afternoon group had a significantly higher proportion of afternoon steps and MVPA than the morning and night groups (Table S5; p < 0.001, both). The night group had a significantly higher proportion of night steps and MVPA than the morning and afternoon groups (Table S5; p < 0.001, both). The characteristics of each group by sex showed a similar trend to that of the overall results (Tables S6 and S7).

### Comparison of diurnal changes in glucose concentrations and AUC

Each group’s 24-hour blood glucose fluctuations by the number of steps are shown in Figure 2 (Figures 2a, all; 2c, male; and 2e, female). The AUC of 24-hour blood glucose fluctuations was calculated and examined, and the results showed that the night group had a significantly lower value than the afternoon group in females (Figure 2f, p < 0.05). However, no significant differences were observed among the groups overall, or in males (Figures 2b and 2d). However, when physical activity patterns were examined using MVPA, no statistically significant differences were found for any of the items (Figure 3).

**Figure 2.**
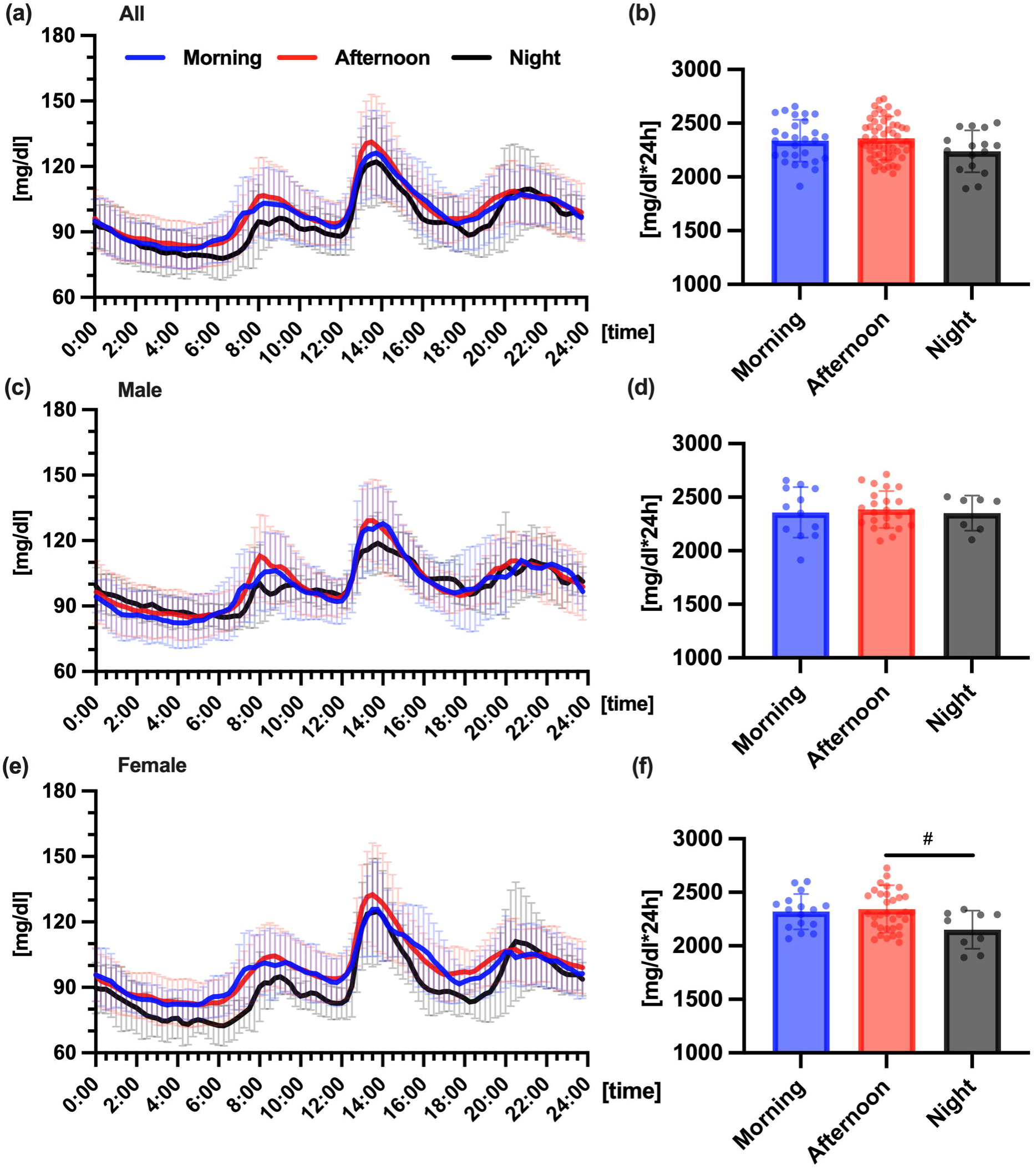
Comparison of diurnal changes in glucose concentrations and area under the curve (AUC), grouped by the number of steps. Comparison of the 24-hour glucose variability (a) and the AUC (b) across groups in all participants, Comparison of 24-hour glucose variability (c) and AUC (d) in males, Comparison of 24-hour blood glucose variability (e) and AUC (f) in females, ^#^ *p* < 0.05 versus afternoon group (One-way ANOVA with Bonferroni). The morning, afternoon, and night groups are represented by blue, red, and dark colors, respectively.

**Figure 3.**
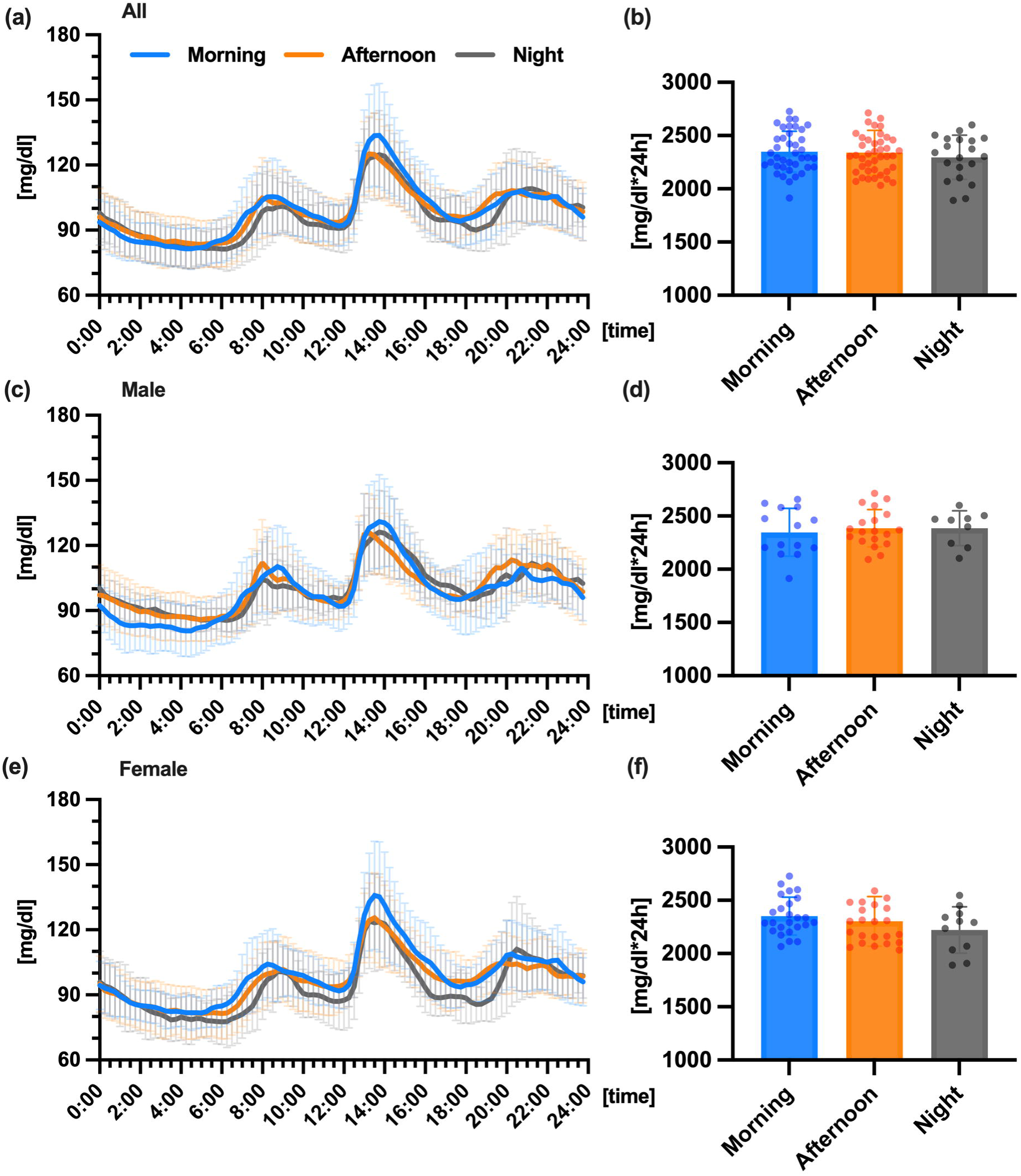
Comparison of diurnal changes in glucose concentrations and area under the curve (AUC), grouped by MVPA. Comparison of the 24-hour glucose variability (a) and AUC (b) across groups in all participants, Comparison of 24-hour glucose variability (c) and AUC (d) in males, Comparison of 24-hour blood glucose variability (e) and AUC (f) in females. The morning, afternoon, and night groups are represented by light blue, orange, and gray colors, respectively.

### Comparison of glucose concentration and AUC after consuming a prescribed meal

When examining physical activity patterns after the prescribed meal using step count, an interaction between time x group was observed in blood glucose fluctuations (p < 0.05), with the night group having significantly lower blood glucose levels 45 minutes after the prescribed meal, compared to the morning and afternoon groups (Figure 4a, p < 0.05), and 60 minutes after the prescribed meal compared to the afternoon group (Figure 4a, p < 0.05). Additionally, the night group showed significantly lower values than the afternoon group when AUC of blood glucose levels were calculated after ingestion of the prescribed meal (Figure 4b, p < 0.05), as well as when age and physical activity levels were included as covariates in the AUC (p < 0.05). However, although similar blood glucose fluctuations were observed when the study was conducted separately for males and females, no statistically significant differences were found among groups (Figures 4c, d, e, and f). Similarly, when physical activity patterns were examined using MVPA, no statistically significant differences were observed among groups in blood glucose fluctuations and AUC after the prescribed meal (Figure 5).

**Figure 4.**
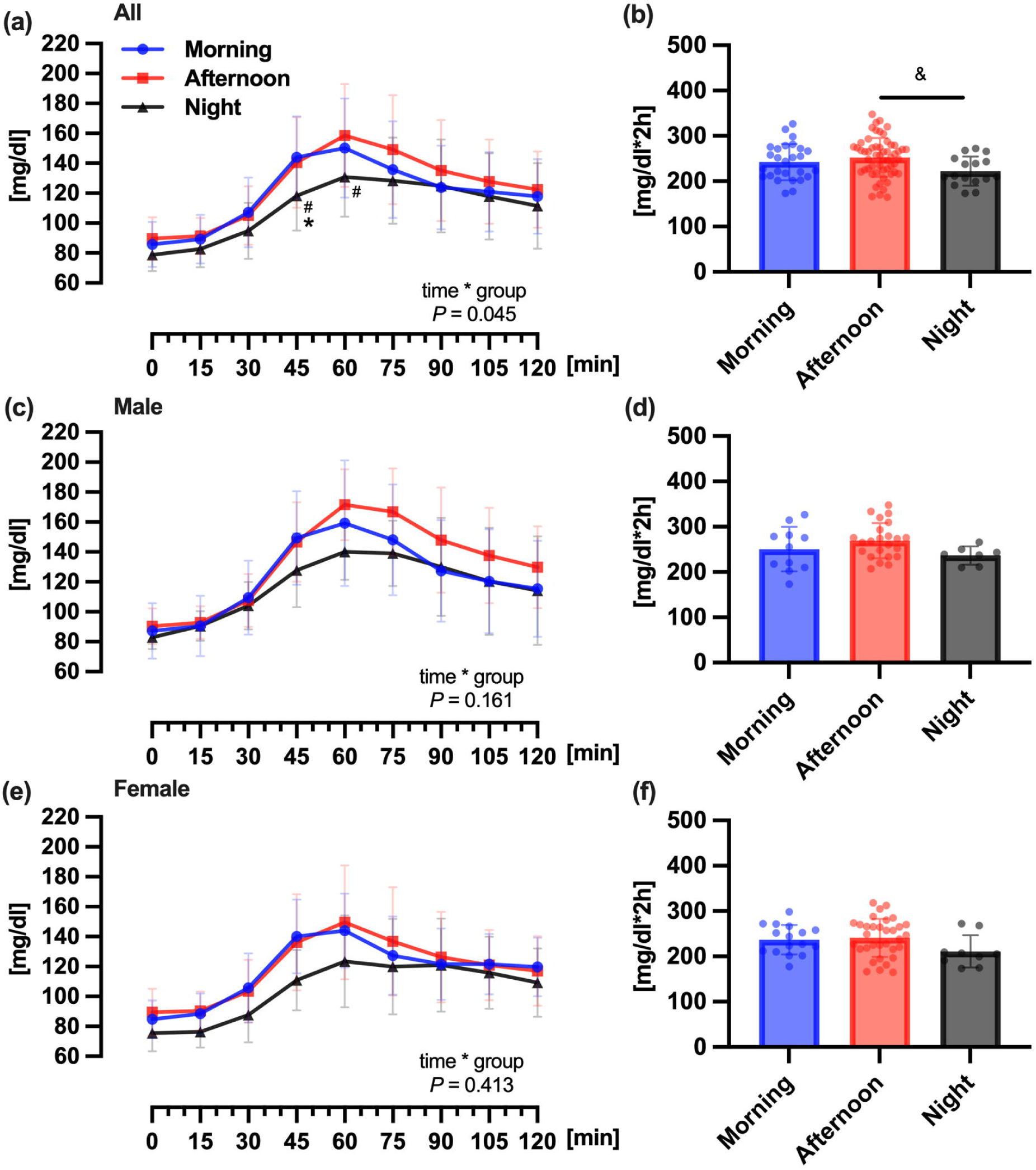
Comparison of glucose concentration and area under the curve (AUC) after consuming a prescribed meal, grouped by the number of steps. Comparison of the glucose concentration (a) and the AUC (b) 2 hours after the prescribed meal consumption across groups in all participants. A prescribed meal was served for breakfast, on the first day (day 1) of the study, * *p* < 0.05 compared with level in the morning group (Bonferroni test for post-hoc), ^#^ *p* < 0.05 compared with level in the afternoon group (Bonferroni test for post-hoc), ^$^ *p* < 0.05 compared with level in the afternoon group (One-way ANOVA with Bonferroni), Comparison of glucose concentration (c) and AUC (d) in males, Comparison of glucose concentration (e) and AUC (f) in females, The morning, afternoon, and night groups are represented by blue, red, and dark colors, respectively.

**Figure 5.**
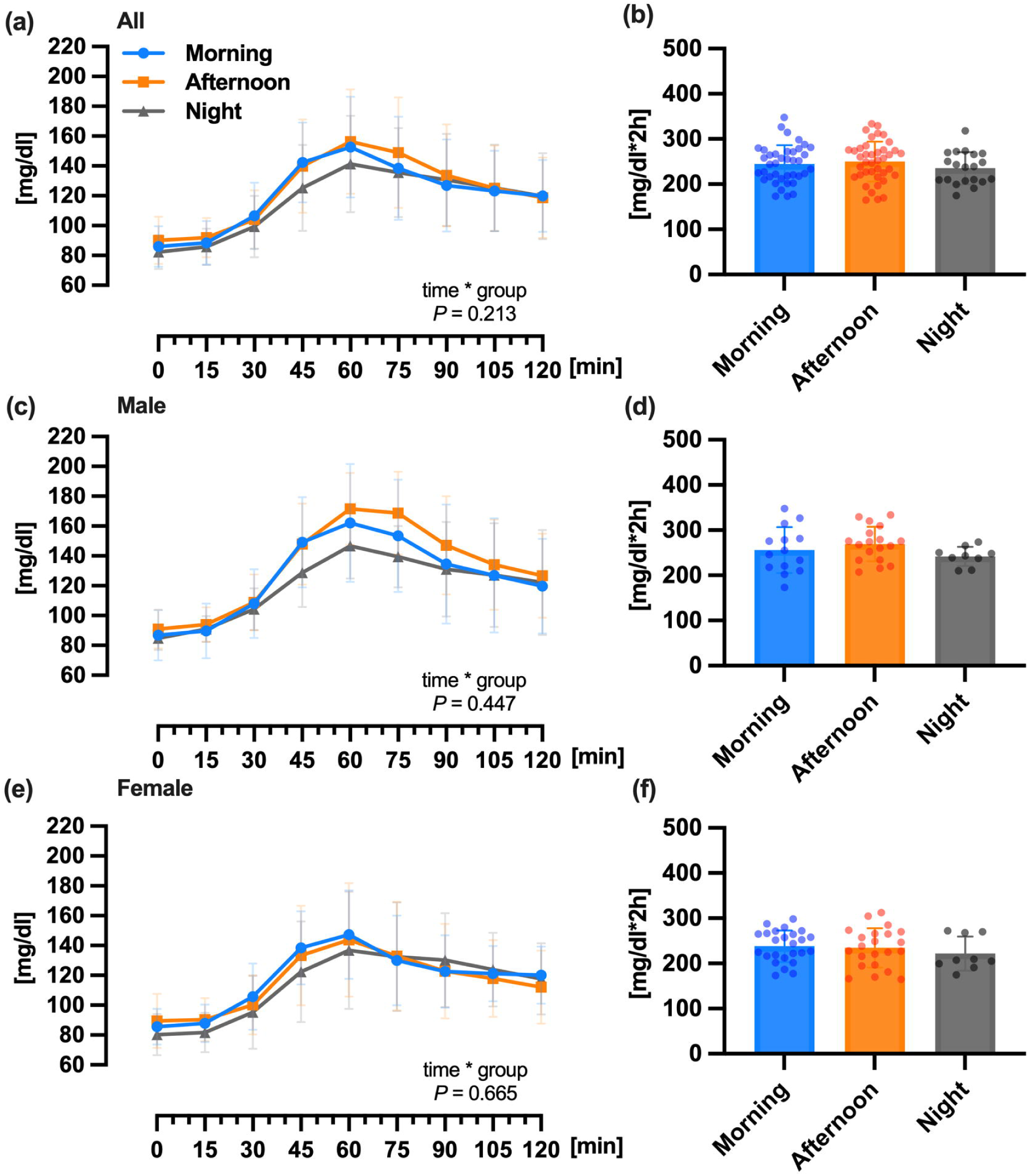
Comparison of glucose concentration and area under the curve (AUC) after consuming a prescribed meal, grouped by intensity. Comparison of glucose concentrations (a) and AUC (b) 2 hours after prescribed meal consumption across groups in all the participants, Comparison of glucose concentration (c) and AUC (d) in males, Comparison of glucose concentration (e) and AUC (f) in females, The morning, afternoon, and night groups are represented by light blue, orange, and gray colors, respectively.

### Comparison of glucose parameters

Several glucose parameters were calculated and examined (Table S8). An examination of physical activity patterns using the number of steps showed, that only females in the night group had significantly lower values of MIN than the afternoon group (p < 0.05, both). The mean was also significantly lower in the night group than in the afternoon group (p < 0.05). However, when physical activity patterns were examined using MVPA, no statistically significant differences were found for any of the parameters among the groups.

## Discussion

This study focused on the relationship between differences in physical activity patterns and blood glucose variability under free-living conditions using CGM. In addition, it used a prescribed diet, that facilitated controlling and studying the effects of diet on blood glucose levels. These results suggest that accumulating evening physical activity in daily life may be effective in controlling blood glucose levels.

This study’s results are consistent with those of previous studies on exercise. A previous study showed that the accumulation of exercise in the afternoon and evening may improve glucose homeostasis ^25^. In addition, the accumulation of evening MVPA is associated with lower blood glucose levels in adults with overweight/obesity and metabolic abnormalities, and this association is reported to be more pronounced in individuals with impaired glucose regulation ^14^. A previous study showed that evening physical activity reduced insulin resistance compared with morning physical activity in overweight or obese individuals in a Dutch cohort ^15^. In addition, previous exercise-based studies have shown that evening physical activity may result in greater benefits for glycemic control compared with morning physical activity ^12,13,26^.

The potential specific mechanisms of daily physical activity patterns on glycemic control remain unclear. The metabolic response to physical activity has been shown to vary depending on the timing ^7–9^: glucose tolerance has been shown to have diurnal variations, and reported to be lower in the evening compared to the morning ^5^. As this has also been associated with reduced insulin sensitivity and β-cell responsiveness, blood glucose levels are more likely to be higher in the evening than in the morning ^6^. In addition to diurnal fluctuations in insulin sensitivity and glucose tolerance, several hormonal factors may also influence the metabolic responses to physical activity at different times of the day. Cortisol levels, which follow a circadian rhythm and typically peak in the early morning, can impact hepatic glucose output and insulin sensitivity ^27,28^. Similarly, melatonin secretion increases in the evening and may impair glucose tolerance and insulin action ^29^. Furthermore, sympathetic nervous system activity and catecholamine levels (e.g., adrenaline) fluctuate throughout the day and may modulate substrate utilization and glucose regulation during physical activity ^30,31^. Sex hormones, including estrogen and progesterone, may also affect glucose metabolism and could contribute to the observed sex-related differences in responses^32^. These hormonal rhythms may partially explain the improved glycemic outcomes observed with evening physical activity in our study. Future research should consider chronobiological and endocrine factors when evaluating the timing of lifestyle interventions. It has also been shown that the efficiency of glucose uptake into skeletal muscles is lower in the evening than in the morning ^25^. Therefore, accumulating physical activity in the evening and later, when insulin resistance is high, may have suppressed the rise in blood glucose levels by promoting substrate utilization, and improving glucose uptake into skeletal muscles.

Most previous studies have focused on MVPA, and examined the relationship between physical activity patterns and blood glucose levels ^14–17^. However, physical activity is important for glycemic control, both, in terms of MVPA and the number of steps. Previous studies have shown an association between the amount of physical activity and blood glucose levels, suggesting that increased physical activity is effective in controlling blood glucose levels in healthy individuals and patients with type 2 diabetes ^18,19^. In this study, the association between physical activity patterns and glycemic variability was more pronounced for the number of steps than MVPA. Therefore, not only MVPA, but also physical activity patterns (number of steps) are important for glucose homeostasis.

Currently, there is no standardized approach to defining diurnal time intervals in physical activity research. However, our primary grouping (06:00–12:00, 12:00–18:00, and 18:00–24:00) was based on prior literature and the average wake-sleep schedules of the Japanese population^33–36^. To validate our classification further, we conducted a sensitivity analysis using alternative time windows (7:00–12:00, 12:00–17:00, and 17:00–24:00). While no statistically significant differences in glycemic variability or postprandial glucose responses were observed between groups under the alternative classification, similar trends were found, particularly an attenuated pattern of glucose fluctuations in the night group (Figure S1–S4). These results suggest that our main findings are not overly sensitive to the specific choice of time window definition.

When stratified by sex, the observed associations between physical activity timing and glycemic outcomes were attenuated and did not reach statistical significance in males, while being weaker in females compared to the overall sample. This may be partly attributable to insufficient statistical power in these subgroup analyses, as the number of participants per sex-stratified group was relatively small. Additionally, potential sex-related differences in hormonal profiles, body composition, and sleep-wake patterns may have influenced glucose metabolism and interacted with the effects of physical activity timing ^32,37^. While our findings suggest a general trend toward lower glycemic variability with evening activity, further studies with larger, sex-balanced samples are needed to confirm sex-specific effects and clarify the underlying mechanisms.

Additionally, while our study focused primarily on physical activity patterns, we recognize that dietary behaviors, particularly meal timing and composition, can also influence glycemic outcomes. To address this, we examined meal timing across physical activity pattern groups (Tables S2–S7). When participants were grouped by step count, only breakfast timing was significantly later in the night group, with no significant differences in lunch or dinner timing. In contrast, no group differences were found when grouping by MVPA. These findings suggest that the overall confounding impact of meal timing on glycemic variability in this study is likely limited. Furthermore, we conducted an ANCOVA adjusting for breakfast timing in the postprandial glucose analysis, where group differences were observed. After adjustment, the difference in postprandial glucose AUC was no longer statistically significant (p = 0.055). Since no group differences in meal timing were observed among females, we did not apply this adjustment in the sex-stratified analyses. Nonetheless, future research should consider more detailed control of dietary variables to further reduce potential confounding.

One major strength of this study is the combined use of triaxial accelerometer data and continuous glucose monitoring (CGM), which enabled a detailed characterization of the relationship between physical activity patterns and glycemic variability under free-living conditions. This methodological integration provides ecologically valid insights that reflect daily life behaviors more accurately than laboratory-based interventions. This study had several limitations. First, as it involved only healthy subjects, it is unclear whether its findings can be applied to people with metabolic diseases, such as diabetes and obesity. Second, the volunteer-based recruitment may have introduced selection bias, as participants were potentially more health-conscious or physically active than the general population, limiting external validity. Third, it collected data of participants, who wore a triaxial accelerometer for just one week, and analyzed their physical activity patterns to examine the relationship with blood glucose fluctuations. Hence, it is unclear whether data on their one-week’s physical activity data would accurately reflect daily life. Therefore, a more detailed study with longer-term data is needed in the future. Fourth, while this study focused on inter-individual differences in habitual activity timing, it did not account for daily fluctuations within individuals. For instance, participants classified in the night group may not have consistently engaged in evening activity each day. Person-day level analyses in future research could help clarify within-person variability and better capture the temporal dynamics of physical activity and glycemic control. Fifth, classifying physical activity relative to meal timing (e.g., postprandial activity window) may offer additional physiological insight and should be explored in future studies. Furthermore, since this study conducted a cross-sectional analysis of the relationship between physical activity pattern and blood glucose fluctuations; therefore, it cannot definitively establish a causal relationship. Finally, each group’s sample sizes relating to physical activity patterns were different. In particular, the night group’s sample size was smaller than that of the morning and afternoon groups. Therefore, future studies need to consider sample sizes.

## Conclusions

This study suggests that the association between daily physical activity patterns and glycemic variability may differ, and that the accumulation of evening physical activity may be more effective for glycemic control. In addition, the association between physical activity patterns and glycemic variability in healthy adults was more pronounced for the number of steps than for MVPA. These findings highlight the need to consider physical activity patterns as a new approach to diabetes prevention in healthy adults.

## Online Methods

### Study participants

This study included healthy adults (n = 107; 62 female and 45 male) aged ≥20 years. It was conducted between June and October 2021. On account of the coronavirus pandemic, mail, web, and social networking services were its primary mode of communication with participants, who were recruited by advertising through posters and flyers. Participants’ inclusion criteria were: (1) Not using glucose/insulin-lowering or related medications; (2) absence of diabetes and thyroid dysfunction; (3) no history of mental illness; (4) not using pacemakers or other implantable medical devices; (5) no history of serious cardiovascular disease, acute cancer, or surgery of the gastrointestinal tract (primarily the esophagus, stomach, duodenum, small intestine, colon, and liver), including appendectomy; (6) no conditions relating to the knee or back, that would interfere with work; (7) no pregnancy; and (8) no regular exercise habits (average: ≥60 minutes/day). Participants were excluded from the study if they did not meet the inclusion criteria (n = 2) or did not wear a triaxial accelerometer (n = 3). Consequently, 102 (60 female and 42 male) participants were included in the analysis (Figure 1).

The protocol for this study conforms to the Declaration of Helsinki. It was approved by the Ethics Committee of the National Institutes of Biomedical Innovation, Health, and Nutrition (approval number: 146). The human trial was registered with the University Hospital Medical Information Network Clinical Trials Registry at: www.umin.ac.jp/ctr/ (number: UMIN000043749). Informed consent was obtained from all the participants after explaining the details of the experiment to them.

### Study design

All participants were fitted with triaxial accelerometers at the waist and glucose monitors to examine the relationship between their physical activity patterns and blood glucose variability under free-living conditions. This study was conducted for 9 days, starting immediately after continuous glucose monitoring (CGM) was assessed (day 0). On the first day of the study (Day 1), a prescribed meal was served for breakfast. Day 2 onwards, the participants wore a triaxial accelerometer to assess their physical activity. In addition, all the participants individually recorded their wake-up time, bedtime, and mealtime using Google Forms at bedtime for one week (between days 2 and 8).

Physical activity data from days two to eight were used to classify participants based on their highest activity level of the day. Since these measures can reflect different aspects of physical behavior, grouping was performed separately using two metrics: the number of steps and the amount of moderate-to-vigorous physical activity (MVPA). For example, step counts may peak in the morning even when MVPA does not. Participants were categorized into morning (06:00–12:00), afternoon (12:00–18:00), or night (18:00–24:00) groups depending on when their step count or MVPA was greatest (Figure 1).

Currently, there is no standardized criterion for defining time intervals when analyzing physical activity patterns. However, several previous studies examining the diurnal variation of physical activity and health outcomes have often categorized activity into three or four time periods. Among these, the time windows of morning (06:00–12:00), afternoon (12:00–18:00), and evening (18:00–24:00) have been frequently used^33–35^. Moreover, according to the “Survey on Time Use and Leisure Activities” conducted by the Statistics Bureau of Japan, the typical wake-up and bedtime for Japanese adults are after 6:00 and after 23:00, respectively^36^. Based on these behavioral patterns, we determined that the 06:00– 24:00 window is appropriate for evaluating daily physical activity accumulation under free-living conditions.

### Test meal

To minimize the influence of dietary habits on blood glucose variability, a prescribed diet for breakfast consumption at home was provided on the first day of the study (day 1). The meal contained 329 kcal, whose energy was derived from 5.3% (17.4 kcal) protein, 30.6% (30.6 kcal) fat, and 64.1% (211 kcal) carbohydrate. Blood glucose levels peak approximately 60 minutes after the start of a meal, and return to pre-meal levels within 2 to 3 hours. However, even if blood glucose levels return to pre-meal levels within 3 hours, absorption of ingested carbohydrates is known to continue for at least 5 to 6 hours after a meal ^38^. Therefore, we minimized the effect on blood glucose levels by eating before the prescribed meal, and using breakfast, which has a longer fasting period, as the prescribed meal.

### Measurements

#### Anthropometry

Body mass was measured to the nearest 0.1 kg, using a digital scale (BC-308; TANITA, Kyoto, Japan). The participants’ sex, age, and height were assessed as baseline information using a self-report questionnaire ^39^. Body mass index (BMI) was calculated as weight (kg) divided by height squared (m).

#### Physical activity assessment

All the participants were asked to wear a triaxial accelerometer (Active Style Pro HJA-750C; Omron Corp., Kyoto, Japan) on their waist for one week (between days 2 and 8). The triaxial accelerometers were worn daily from morning to night, except during bathing and sleeping. The device determined the level of intensity through metabolic equivalents (METs) generated by the activity every 1 minute, using a Classification Algorithm for ambulatory and non-ambulatory activities, which leads to an accurate prediction of METs, even for non-ambulatory activities ^40,41^.

Data were included from participants who wore the accelerometer for at least 10 hours (600 minutes) per day on at least five weekdays and one weekend day. Step count and MVPA were assessed. Minutes ≥ 3 METs were classified as MVPA ^42^. A macro program developed and distributed by the Japan Physical Activity Research Platform was used for processing and calculating the accelerometer data ^43^, which was thereafter grouped using a Python code.

#### Glucose variability

All the participants were required to wear a CGM system (FreeStyle Libre; Abbott Laboratories, Chicago, Illinois, USA) to continuously measure blood glucose levels during the study period (between days 0 and 8). The sensor was placed on the back of the upper arm. Blood glucose levels were assessed under free-living conditions (7 days), and data were collected every 15 minutes using CGM. The CGM used in this study required data to be read every 8 hours, or data would be missed. Therefore, the analysis was performed on days when at least 75% of the per day data could be confirmed ^44^. The 24-hour blood glucose variability and AUC were calculated for each group. In addition, blood glucose levels and AUC were calculated up to 2 hours after the prescribed meal. A glycemic variability calculator (a macro-enabled Excel workbook, EasyGV, University of Oxford, UK) was used to evaluate mean glucose level, standard deviation, continuous overall net glycemic action, J-index, mean amplitude of glycemic excursion, maximum glucose level, and minimum glucose level (MIN). The CGM data were time-matched and analyzed with the accelerometer data.

#### Dietary assessment

The participants’ energy intake was assessed using a brief-type self-administered diet history questionnaire, which is a relatively simple questionnaire designed to assess the amount of nutrients habitually consumed from regular foods (excluding supplements, etc.) on an individual basis, and provide nutrient and food intakes for each person. It consists of approximately 80 questions, and calculates intakes of 58 foods and more than 100 nutrients ^45,46^. With self-estimated dietary assessments, avoiding under or overestimation of dietary intake is not easy ^47^. Therefore, nutrient intakes were estimated by adjusting the reported dietary intake based on each participant’s energy expenditure calculated, using the accelerometer.

#### Statistical analysis

All measurements are presented as mean ± standard deviation. The Shapiro-Wilk test was used to analyze normal or non-normal distributions prior to statistical processing. Unpaired Student’s *t*-tests were used to compare the physical characteristics of male and female participants. One-way analysis of variance (ANOVA) was used to compare the AUC, physical activity, sleep and meal times, energy intake, and glycemic variability among the groups. One-way ANOVA with Dunnett’s correction was used for comparisons when equal variances were not assumed. Two-way ANOVA was used to compare glucose variability among groups after the prescribed meal intake, and determine the effects of time and group, as factors. When a significant interaction was observed, the Bonferroni method was used for post hoc comparisons. In addition, analyses were performed using analysis of covariance with age, meal time or physical activity levels (number of steps or MVPA) as a covariate. To assess the robustness of the findings, sensitivity analyses were conducted using alternative time windows for defining time-of-day groups (i.e., morning: 07:00–12:00, afternoon: 12:00–17:00, and night: 17:00–24:00), and the outcomes were re-evaluated accordingly. IBM SPSS Statistics for Mac (version 28.0; IBM Corp., Armonk, NY, USA) was used for data analysis with statistical significance thresholds. Statistical significance was set at *P* < 0.05.

## Data availability statement

The data supporting the results of this study are available upon request from the corresponding author, Eiichi Yoshimura. The data are not publicly available due to restrictions that include information that could compromise the privacy of research participants.

## Acknowledgments

The authors thank the participants and collaborators associated with this study.

## Author contributions

Hyeon-Ki Kim, Eiichi Yoshimura, and Motohiko Miyachi: Conceptualization. Hyeon-Ki Kim, Eiichi Yoshimura, Yuka Hamada, Yi-An Chen, and Yoichi Hatamoto: Data curation. Hyeon-Ki Kim, Eiichi Yoshimura, and Yoichi Hatamoto: Formal analysis. Eiichi Yoshimura, Yuka Hamada, Yoichi Hatamoto, Hinako Nanri, Motohiko Miyachi, and Takashi Nakagata: Investigation. Hyeon-Ki Kim and Eiichi Yoshimura: Methodology. Eiichi Yoshimura and Motohiko Miyachi: Project administration. Eiichi Yoshimura, Yu Tahara, and Shigenobu Shibata: Resources. Eiichi Yoshimura and Motohiko Miyachi: Supervision. Hyeon-Ki Kim: Visualization. Hyeon-Ki Kim and Eiichi Yoshimura: Writing—original draft. Yuka Hamada, Yoichi Hatamoto, Mana Hatanaka, Hinako Nanri, Takashi Nakagata, Naoyuki Matsumoto, Seiya Shimoda, Shigeho Tanaka, Motohiko Miyachi, Yi-An Chen, Yu Tahara, and Shigenobu Shibata: Writing—review & editing. All the authors have read and approved the final manuscript.

## Ethics declarations

Competing Interests

All authors declare no other competing interests.

