## Supplementary Figures for "Relationship between differences in physical activity patterns and blood glucose variability under free-living conditions in adults without diabetes"

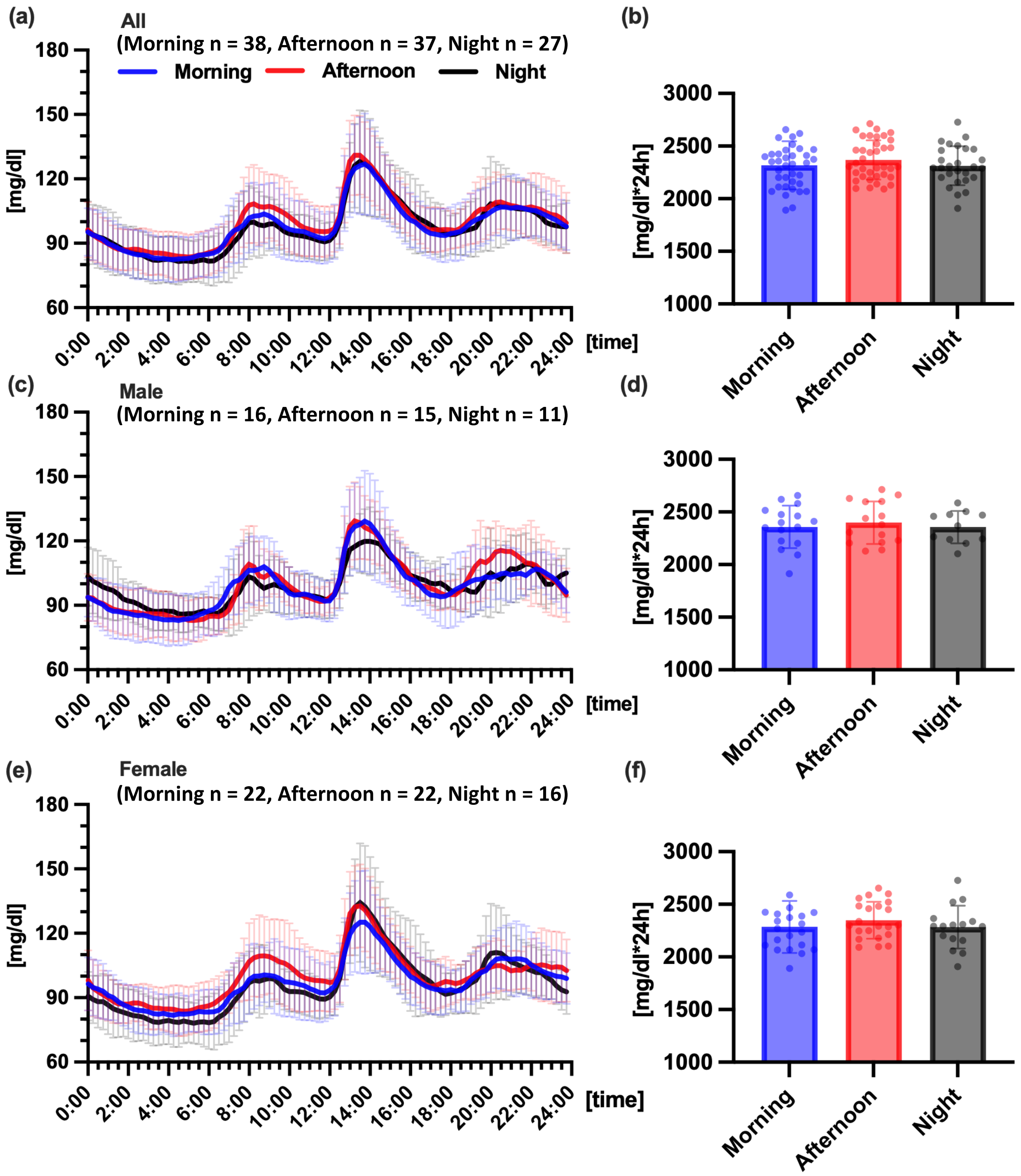


**Supplementary Figure 1 Comparison of diurnal changes in glucose concentrations and area under the curve (AUC), grouped by the number of steps.** Comparison of the 24-hour glucose variability (a) and the AUC (b) across groups in all participants, Comparison of 24-hour glucose variability (c) and AUC (d) in males, Comparison of 24-hour blood glucose variability (e) and AUC (f) in females. The morning, afternoon, and night groups are represented by blue, red, and dark colors, respectively.

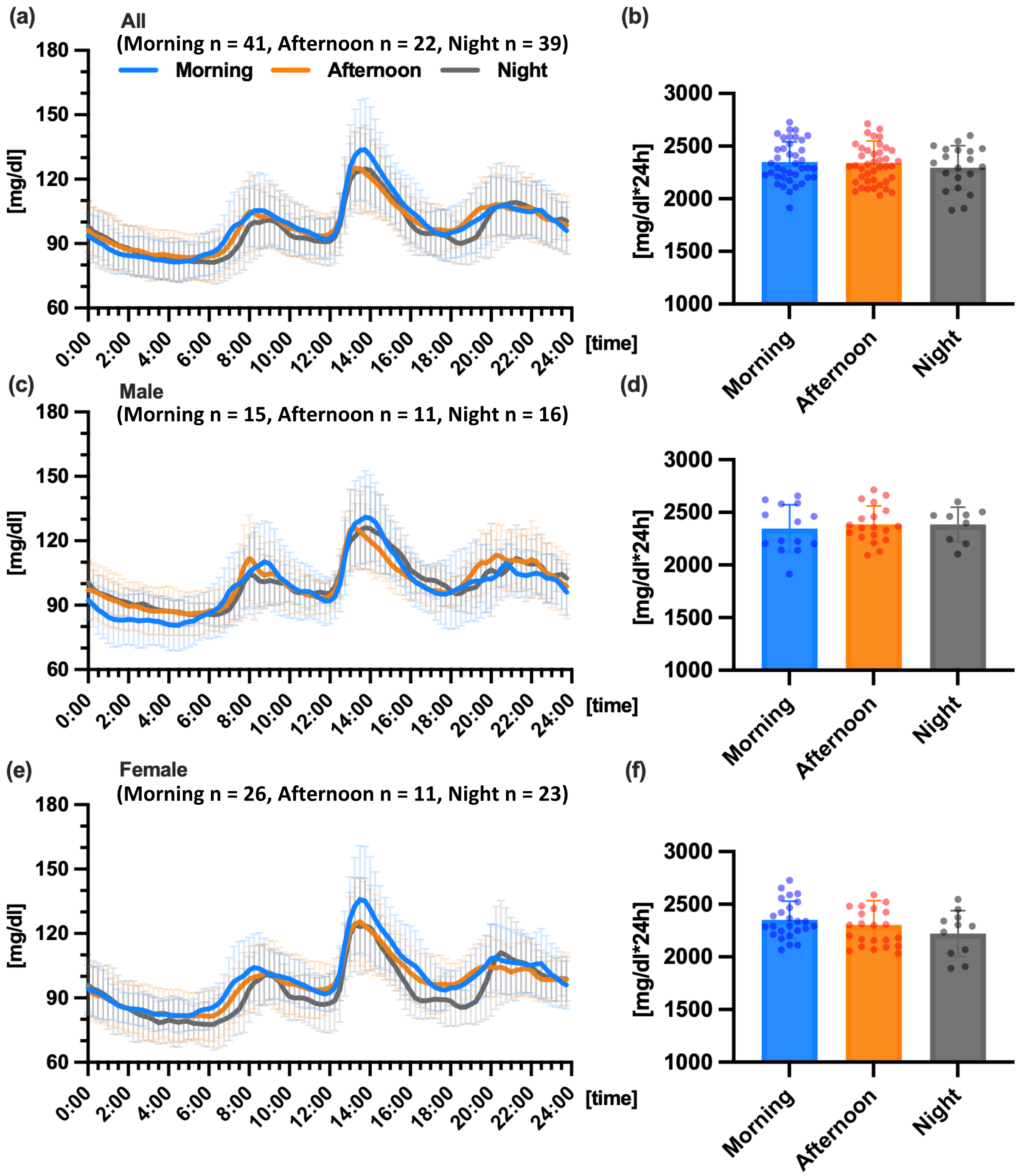


**Supplementary Figure 2 Comparison of diurnal changes in glucose concentrations and area under the curve (AUC), grouped by MVPA.** Comparison of the 24-hour glucose variability (a) and AUC (b) across groups in all participants, Comparison of 24-hour glucose variability (c) and AUC (d) in males, Comparison of 24-hour blood glucose variability (e) and AUC (f) in females. The morning, afternoon, and night groups are represented by light blue, orange, and gray colors, respectively.

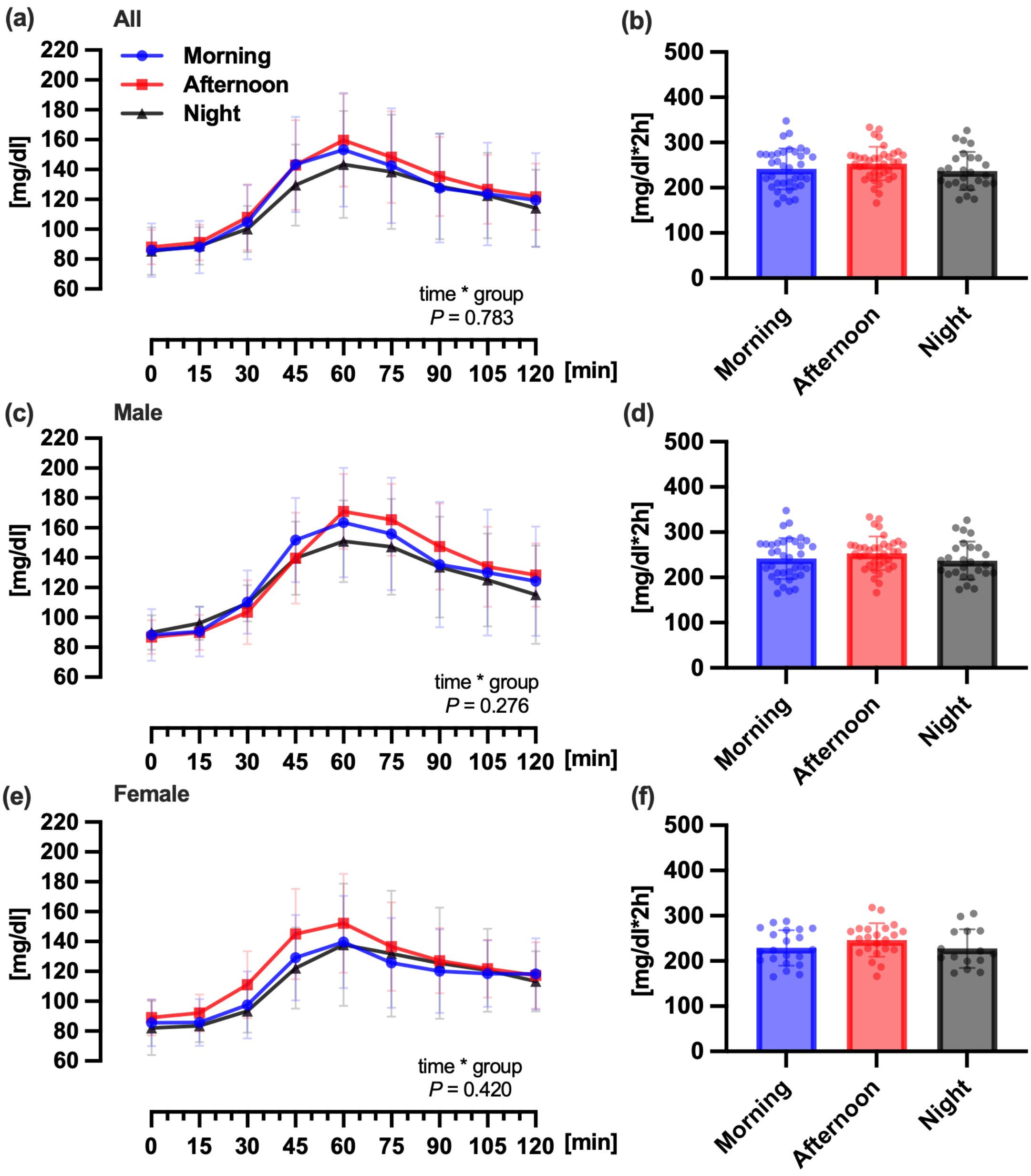


**Supplementary Figure 3 Comparison of glucose concentration and area under the curve (AUC) after consuming a prescribed meal, grouped by the number of steps.** Comparison of the glucose concentration (a) and the AUC (b) 2 hours after the prescribed meal consumption across groups in all participants. A prescribed meal was served for breakfast, on the first day (day 1) of the study. Comparison of glucose concentration (c) and AUC (d) in males, Comparison of glucose concentration (e) and AUC (f) in females. The morning, afternoon, and night groups are represented by blue, red, and dark colors, respectively.

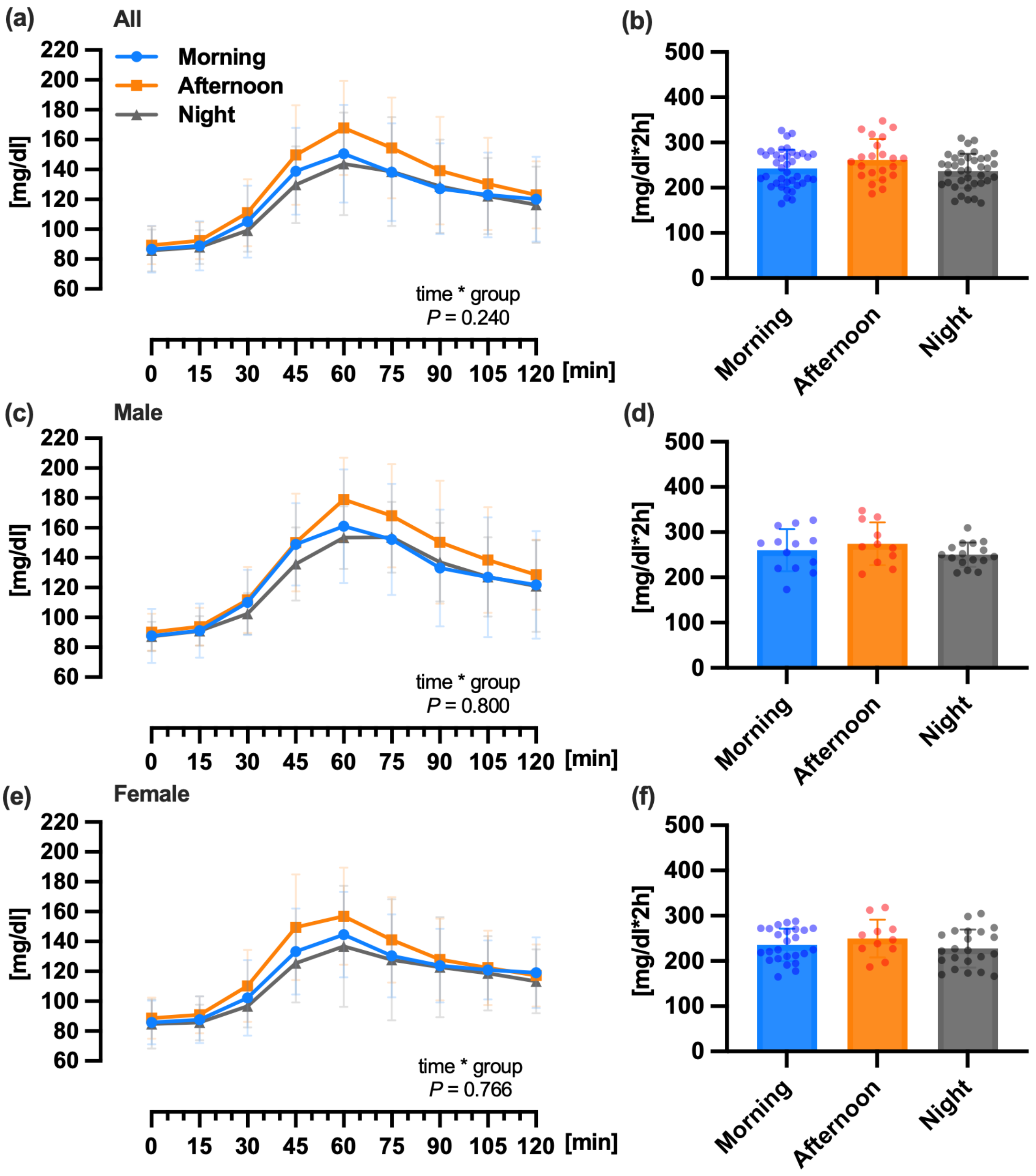


**Supplementary Figure 4 Comparison of glucose concentration and area under the curve (AUC) after consuming a prescribed meal, grouped by intensity.** Comparison of glucose concentrations (a) and AUC (b) 2 hours after prescribed meal consumption across groups in all the participants, Comparison of glucose concentration (c) and AUC (d) in males, Comparison of glucose concentration (e) and AUC (f) in females, The morning, afternoon, and night groups are represented by light blue, orange, and gray colors, respectively.
