## Supplementary Tables for "Relationship between differences in physical activity patterns and blood glucose variability under free-living conditions in adults without diabetes"

Supplementary table 1. Physical characteristics of the participants.

| | All (n = 102) | Male (n = 42) | Female (n = 60) | *P*-value |
|---|---|---|---|---|
| **Physical characteristics** | | | | |
| Age (years) | 39.8 ± 11.8 | 40.4 ± 11.7 | 39.4 ± 12.1 | 0.674 |
| Height (cm) | 165.4 ± 7.8 | 171.9 ± 5.4 | 160.6 ± 5.2 | **< 0.001** |
| Weight (kg) | 62.5 ± 12.8 | 70.3 ± 12.5 | 56.5 ± 9.6 | **< 0.001** |
| BMI (kg/m2) | 22.8 ± 4.0 | 23.7 ± 3.8 | 21.9 ± 3.9 | **0.023** |
| **Physical activity level** | | | | |
| Total step count (steps/day) | 7050.2 ± 2893.2 | 7265.5 ± 3695.3 | 6901.8 ± 2200.9 | 0.570 |
| Morning (steps/day) | 2324.7 ± 1253.9 | 2391.7 ± 1684.5 | 2278.5 ± 853.5 | 0.655 |
| Afternoon (steps/day) | 2708.3 ± 988.4 | 2810.3 ± 1190.9 | 2638.1 ± 824.2 | 0.419 |
| Night (steps/day) | 1915.3 ± 1502.2 | 1947.9 ± 1747.9 | 1892.9 ± 1321.8 | 0.856 |
| Morning steps/total steps (%) | 33.4 ± 9.0 | 33.4 ± 10.2 | 33.5 ± 8.2 | 0.937 |
| Afternoon steps/total steps (%) | 39.3 ± 9.1 | 39.6 ± 9.3 | 39.2 ± 9.0 | 0.819 |
| Night steps/total steps (%) | 25.7 ± 11.4 | 25.3 ± 12.4 | 26.0 ± 10.7 | 0.763 |
| Total MVPA (min/day) | 53.0 ± 25.5 | 52.8 ± 30.0 | 53.1 ± 22.1 | 0.763 |
| Morning MVPA (min/day) | 18.0 ± 10.2 | 17.3 ± 13.0 | 18.4 ± 7.8 | 0.941 |
| Afternoon MVPA (min/day) | 18.6 ± 9.4 | 19.2 ± 11.0 | 18.3 ± 8.2 | 0.654 |
| Night MVPA (min/day) | 15.3 ± 12.6 | 15.1 ± 14.0 | 15.5 ± 11.6 | 0.880 |
| Morning MVPA/total MVPA (%) | 34.8 ± 10.4 | 33.3 ± 12.0 | 35.8 ± 9.1 | 0.254 |
| Afternoon MVPA/total MVPA (%) | 36.1 ± 10.0 | 37.6 ± 10.9 | 35.1 ± 9.2 | 0.224 |
| Night MVPA/total MVPA (%) | 27.2 ± 12.3 | 27.0 ± 13.9 | 27.3 ± 11.2 | 0.904 |
| **Sleep and meal time** | | | | |
| Wake-up time (hh:mm) | 6:46 ± 1:02 | 6:49 ± 1:19 | 6:44 ± 0:47 | 0.725 |
| Bedtime (hh:mm) | 23:59 ± 1:07 | 0:06 ± 1:14 | 23:53 ± 1:01 | 0.411 |
| Breakfast (hh:mm) | 7:36 ± 0:45 | 7:28 ± 0:47 | 7:43 ± 0:42 | 0.130 |
| Lunch (hh:mm) | 12:31 ± 0:29 | 12:29 ± 0:28 | 12:33 ± 0:30 | 0.362 |
| Dinner (hh:mm) | 19:47 ± 1:09 | 19:39 ± 1:23 | 19:52 ± 0:57 | 0.936 |
| **Dietary intake** | | | | |
| Total energy intake (kcal/day) | 2210.7 ± 377.0 | 2518.0 ± 332.8 | 2004.2 ± 239.9 | **< 0.001** |
| Protein (g/day) | 87.7 ± 19.6 | 99.3 ± 20.4 | 79.9 ± 14.7 | **< 0.001** |
| Fat (g/day) | 76.5 ± 19.0 | 86.6 ± 20.6 | 69.8 ± 14.4 | **< 0.001** |
| Carbohydrate (g/day) | 262.1 ± 61.6 | 294.5 ± 70.8 | 240.3 ± 42.9 | **< 0.001** |
| Dietary fiber (g/day) | 13.9 ± 4.4 | 15.4 ± 5.3 | 12.8 ± 3.3 | **0.009** |

All data are presented as the mean ± standard deviation. *P*-values in bold indicate significant differences between male and female as calculated by Student's t-test. BMI: body mass index, MVPA: moderate-to-vigorous physical activity.

Supplementary table 2. Physical characteristics of participants by grouping according to step count.

| All (n = 102) | Grouping by number of steps | | |
|---|---|---|---|
| | **Morning (n = 29)** | **Afternoon (n = 57)** | **Night (n = 16)** |
| **Physical characteristics** | | | |
| Age (years) | 44.9 ± 11.3 | 38.4 ± 10.7 * | 35.9 ± 14.5 * |
| Height (cm) | 164.7 ± 6.6 | 165.6 ± 8.2 | 164.8 ± 8.0 |
| Weight (kg) | 62.7 ± 14.6 | 62.4 ± 12.8 | 60.2 ± 9.7 |
| BMI (kg/m2) | 22.9 ± 4.0 | 22.7 ± 4.2 | 22.1 ± 3.3 |
| **Physical activity level** | | | |
| Total step count (steps/day) | 6974.1 ± 3693.7 | 6576.9 ± 1996.3 | 8903.5 ± 3430.3 |
| Morning (steps/day) | 3102.7 ± 1946.8 | 2014.0 ± 623.4 # | 2040.7 ± 713.8 # |
| Afternoon (steps/day) | 2574.2 ± 1282.6 | 2843.4 ± 837.0 | 2461.6 ± 860.0 |
| Night (steps/day) | 1372.9 ± 808.5 | 1571.6 ± 890.3 | 4144.2 ± 2188.2 #$ |
| Morning steps/total steps (%) | 43.7 ± 5.7 | 30.9 ± 5.8 *** | 24.1 ± 6.6 ***&&& |
| Afternoon steps/total steps (%) | 33.4 ± 4.6 | 45.2 ± 6.6 *** | 28.7 ± 5.7 *&&& |
| Night steps/total steps (%) | 20.9 ± 8.5 | 22.8 ± 7.3 *** | 45.1 ± 8.1 ***&&& |
| Total MVPA (min/day) | 49.3 ± 27.9 | 50.4 ± 23.0 | 69.0 ± 25.4 *& |
| Morning MVPA (min/day) | 22.6 ± 14.3 | 16.2 ± 7.8 * | 15.9 ± 6.1 |
| Afternoon MVPA (min/day) | 15.3 ± 9.8 | 20.3 ± 9.0 | 18.6 ± 8.8 |
| Night MVPA (min/day) | 10.1 ± 7.1 | 13.0 ± 8.7 | 33.1 ± 16.8 #$ |
| Morning MVPA/total MVPA (%) | 45.8 ± 6.0 | 32.1 ± 7.6 *** | 24.6 ± 8.5 ***&&& |
| Afternoon MVPA/total MVPA (%) | 30.3 ± 6.8 | 41.6 ± 8.4 *** | 26.6 ± 6.3 &&& |
| Night MVPA/total MVPA (%) | 21.5 ± 10.2 | 24.6 ± 8.4 | 46.7 ± 9.1 ***&&& |
| **Sleep and meal time** | | | |
| Wake-up time (hh:mm) | 6:12 ± 0:48 | 6:47 ± 0:56 | 7:38 ± 1:12 |
| Bedtime (hh:mm) | 23:30 ± 1:03 | 23:56 ± 1:03 | 0:53 ± 0:56 ***&& |
| Breakfast (hh:mm) | 7:20 ± 0:45 | 7:37 ± 0:42 | 8:02 ± 0:43 ** |
| Lunch (hh:mm) | 12:30 ± 0:29 | 12:29 ± 0:28 | 12:38 ± 0:28 |
| Dinner (hh:mm) | 19:44 ± 0:49 | 19:54 ± 1:03 | 20:02 ± 0:44 |
| **Dietary intake** | | | |
| Total energy intake (kcal/day) | 2185.9 ± 370.1 | 2223.9 ± 383.2 | 2206.4 ± 388.2 |
| Protein (g/day) | 90.4 ± 21.7 | 86.9 ± 18.0 | 85.7 ± 22.3 |
| Fat (g/day) | 77.3 ± 17.8 | 76.0 ± 19.2 | 77.1 ± 21.5 |
| Carbohydrate (g/day) | 265.1 ± 60.6 | 263.6 ± 63.9 | 251.4 ± 57.0 |
| Dietary fiber (g/day) | 14.6 ± 5.1 | 13.9 ± 4.0 | 12.6 ± 4.3 |

All data are presented as the mean ± standard deviation. BMI: body mass index, MVPA: moderate-to-vigorous physical activity. *** $p < 0.001$, ** $p < 0.01$, * $p < 0.05$ vs morning (One-way ANOVA with Bonferroni), &&& $p < 0.001$, && $p < 0.01$, & $p < 0.05$ vs afternoon (One-way ANOVA with Bonferroni), # $p < 0.05$ vs morning (One-way ANOVA with Dunnett), $ $p < 0.05$ vs afternoon (One-way ANOVA with Dunnett).

Supplementary table 3. Physical characteristics of participants by grouping according to step count in males.

| Male (n = 42) | Grouping by number of steps | | |
|---|---|---|---|
| | **Morning (n = 12)** | **Afternoon (n = 23)** | **Night (n = 7)** |
| **Physical characteristics** | | | |
| Age (years) | 46.1 ± 10.8 | 40.4 ± 10.7 | 30.7 ± 11.6 * |
| Height (cm) | 169.9 ± 5.4 | 172.9 ± 5.8 | 172.2 ± 3.0 |
| Weight (kg) | 75.3 ± 12.1 | 69.2 ± 13.4 | 65.3 ± 7.5 |
| BMI (kg/m2) | 26.0 ± 3.1 | 23.1 ± 4.2 | 22.0 ± 2.0 |
| **Physical activity level** | | | |
| Total step count (steps/day) | 7379.3 ± 5084.4 | 6404.3 ± 2156.9 | 9900.5 ± 4229.9 |
| Morning (steps/day) | 3441.0 ± 2752.7 | 1956.7 ± 685.1 | 2022.1 ± 830.4 |
| Afternoon (steps/day) | 3068.4 ± 1641.3 | 2670.2 ± 952.2 | 2828.1 ± 1103.9 |
| Night (steps/day) | 1194.9 ± 662.4 | 1499.7 ± 982.5 | 4711.2 ± 2391.7 $^{\#\$}$ |
| Morning steps/total steps (%) | 45.3 ± 3.8 | 30.7 ± 7.1 *** | 21.7 ± 5.9 ***$^{\&\&\&}$ |
| Afternoon steps/total steps (%) | 33.7 ± 4.4 | 45.8 ± 7.3 *** | 29.3 ± 5.1 $^{\&\&\&}$ |
| Night steps/total steps (%) | 18.7 ± 7.4 | 22.2 ± 8.2 *** | 47.1 ± 5.7 $^{\&\&\&}$ |
| Total MVPA (min/day) | 51.1 ± 36.0 | 46.6 ± 24.2 * | 75.7 ± 29.6 |
| Morning MVPA (min/day) | 24.9 ± 19.7 | 13.9 ± 7.9 | 15.5 ± 7.0 |
| Afternoon MVPA (min/day) | 16.3 ± 12.6 | 19.7 ± 10.1 | 22.2 ± 11.1 |
| Night MVPA (min/day) | 8.0 ± 6.0 | 12.2 ± 9.0 | 36.8 ± 16.9 $^{\#\#\#\$\$\$}$ |
| Morning MVPA/total MVPA (%) | 47.8 ± 4.9 | 29.3 ± 8.3 *** | 21.9 ± 7.6 ***$^{\&}$ |
| Afternoon MVPA/total MVPA (%) | 31.5 ± 7.9 | 43.6 ± 9.5 *** | 27.9 ± 6.4 $^{\&\&\&}$ |
| Night MVPA/total MVPA (%) | 18.4 ± 10.5 | 25.0 ± 10.4 | 48.2 ± 6.5 ***$^{\&\&\&}$ |
| **Sleep and meal time** | | | |
| Wake-up time (hh:mm) | 6:13 ± 0:39 | 6:43 ± 1:15 | 8:07 ± 1:42 **$^{\&}$ |
| Bedtime (hh:mm) | 23:37 ± 1:01 | 23:58 ± 1:14 | 1:12 ± 0:58 *$^{\&}$ |
| Breakfast (hh:mm) | 7:06 ± 0:38 | 7:27 ± 0:46 | 8:10 ± 0:48 * |
| Lunch (hh:mm) | 12:22 ± 0:22 | 12:25 ± 0:24 | 12:47 ± 0:36 |
| Dinner (hh:mm) | 19:45 ± 0:38 | 19:55 ± 1:09 | 20:03 ± 0:38 |
| **Dietary intake** | | | |
| Total energy intake (kcal/day) | 2554.2 ± 225.2 | 2501.8 ± 397.6 | 2514.5 ± 265.9 |
| Protein (g/day) | 105.3 ± 22.6 | 97.2 ± 18.6 | 96.8 ± 23.8 |
| Fat (g/day) | 89.2 ± 13.8 | 85.0 ± 22.9 | 87.7 ± 24.0 |
| Carbohydrate (g/day) | 312.5 ± 56.3 | 291.1 ± 77.0 | 277.5 ± 73.3 |
| Dietary fiber (g/day) | 16.7 ± 6.9 | 15.9 ± 4.5 | 11.6 ± 3.7 |

All data are presented as the mean ± standard deviation. BMI: body mass index, MVPA: moderate-to-vigorous physical activity. *** $p < 0.001$, ** $p < 0.01$, * $p < 0.05$ vs morning (One-way ANOVA with Bonferroni), $^{\&\&\&}$ $p < 0.001$, $^{\&}$ $p < 0.05$ vs afternoon (One-way ANOVA with Bonferroni), $^{\#}$ $p < 0.05$ vs morning (One-way ANOVA with Dunnett), $^{\$}$ $p < 0.05$ vs afternoon (One-way ANOVA with Dunnett).

Supplementary table 4. Physical characteristics of participants by grouping according to step count in females.

| Female (n = 60) | Grouping by number of steps | | |
|---|---|---|---|
| | Morning (n = 17) | Afternoon (n = 34) | Night (n = 9) |
| **Physical characteristics** | | | |
| Age (years) | 44.1 ± 11.8 | 36.9 ± 10.7 | 39.9 ± 15.8 |
| Height (cm) | 161.1 ± 4.8 | 160.7 ± 5.3 | 159.1 ± 5.4 |
| Weight (kg) | 53.8 ± 8.3 | 57.8 ± 10.2 | 56.3 ± 9.8 |
| BMI (kg/m2) | 20.7 ± 3.0 | 22.4 ± 4.2 | 22.3 ± 4.1 |
| **Physical activity level** | | | |
| Total step count (steps/day) | 6688.1 ± 2427.5 | 6690.4 ± 1907.3 | 8128.0 ± 2664.3 |
| Morning (steps/day) | 2863.9 ± 1129.9 | 2051.7 ± 586.6 $^{\#}$ | 2055.1 ± 661.5 |
| Afternoon (steps/day) | 2225.4 ± 845.3 | 2957.2 ± 744.4 ** | 2176.5 ± 513.7 $^{\&}$ |
| Night (steps/day) | 1498.5 ± 895.4 | 1618.9 ± 835.6 | 3703.2 ± 2046.3 $^{\#\$}$ |
| Morning steps/total steps (%) | 42.7 ± 6.6 | 31.0 ± 4.8 *** | 26.1 ± 6.9 ***$^{\&\&\&}$ |
| Afternoon steps/total steps (%) | 33.2 ± 4.8 | 44.9 ± 6.2 *** | 28.3 ± 6.4 $^{\$\$\$}$ |
| Night steps/total steps (%) | 22.4 ± 9.0 | 23.3 ± 6.7 | 43.5 ± 9.7 ***$^{\&\&\&}$ |
| Total MVPA (min/day) | 48.0 ± 21.6 | 52.9 ± 22.1 | 63.9 ± 22.0 |
| Morning MVPA (min/day) | 21.0 ± 9.2 | 17.7 ± 7.4 | 16.3 ± 5.8 |
| Afternoon MVPA (min/day) | 14.6 ± 7.5 | 20.7 ± 8.3 * | 15.8 ± 5.7 |
| Night MVPA (min/day) | 11.5 ± 7.5 | 13.6 ± 8.6 | 30.2 ± 17.1 $^{\#}$ |
| Morning MVPA/total MVPA (%) | 44.5 ± 6.4 | 34.0 ± 6.6 *** | 26.7 ± 9.0 ***$^{\&\&\&}$ |
| Afternoon MVPA/total MVPA (%) | 29.4 ± 6.0 | 40.3 ± 7.5 *** | 25.6 ± 6.4 $^{\&\&\&}$ |
| Night MVPA/total MVPA (%) | 23.8 ± 9.6 | 24.3 ± 6.9 | 45.6 ± 10.9 ***$^{\&\&\&}$ |
| **Sleep and meal time** | | | |
| Wake-up time (hh:mm) | 6:11 ± 0:56 | 6:50 ± 0:39 * | 7:15 ± 0:24 ** |
| Bedtime (hh:mm) | 23:25 ± 1:06 | 23:54 ± 0:55 | 0:38 ± 0:53 * |
| Breakfast (hh:mm) | 7:31 ± 0:48 | 7:44 ± 0:39 | 7:56 ± 0:41 |
| Lunch (hh:mm) | 12:36 ± 0:34 | 12:32 ± 0:31 | 12:31 ± 0:19 |
| Dinner (hh:mm) | 19:44 ± 0:58 | 19:54 ± 0:59 | 20:02 ± 0:50 |
| **Dietary intake** | | | |
| Total energy intake (kcal/day) | 1947.6 ± 214.0 | 2041.3 ± 239.6 | 1966.8 ± 286.1 |
| Protein (g/day) | 80.7 ± 14.9 | 80.2 ± 14.2 | 77.0 ± 17.6 |
| Fat (g/day) | 69.7 ± 16.0 | 70.0 ± 13.6 | 68.9 ± 16.2 |
| Carbohydrate (g/day) | 234.4 ± 40.8 | 245.5 ± 46.5 | 231.1 ± 31.6 |
| Dietary fiber (g/day) | 13.2 ± 2.9 | 12.5 ± 3.1 | 13.3 ± 4.8 |

All data are presented as the mean ± standard deviation. BMI: body mass index, MVPA: moderate-to-vigorous physical activity. *** $p < 0.001$, ** $p < 0.01$, * $p < 0.05$ vs morning (One-way ANOVA with Bonferroni), $^{\&\&\&}$ $p < 0.001$, $^{\&}$ $p < 0.05$ vs afternoon (One-way ANOVA with Bonferroni), $^{\#}$ $p < 0.05$ vs morning (One-way ANOVA with Dunnett), $^{\$}$ $p < 0.05$ vs afternoon (One-way ANOVA with Dunnett).

Supplementary table 5. Physical characteristics of participants by grouping according to the MVPA.

| All (n = 102) | Grouping by MVPA | | |
|---|---|---|---|
| | **Morning (n = 40)** | **Afternoon (n = 42)** | **Night (n = 20)** |
| **Physical characteristics** | | | |
| Age (years) | 45.3 ± 10.9 | 37.2 ± 10.1 ** | 34.6 ± 13.4 ** |
| Height (cm) | 163.8 ± 6.7 | 166.2 ± 8.3 | 166.0 ± 8.2 |
| Weight (kg) | 60.5 ± 12.9 | 64.4 ± 14.2 | 60.7 ± 8.6 |
| BMI (kg/m2) | 22.4 ± 3.6 | 23.3 ± 4.7 | 22.0 ± 3.0 |
| **Physical activity level** | | | |
| Total step count (steps/day) | 7078.1 ± 3139.9 | 6369.6 ± 2141.9 | 8422.1 ± 3357.7 & |
| Morning (steps/day) | 2951.1 ± 1667.3 | 1866.9 ± 577.1 ### | 2001.8 ± 670.1 ### |
| Afternoon (steps/day) | 2653.4 ± 1114.4 | 2865.9 ± 903.6 | 2426.9 ± 904.0 |
| Night (steps/day) | 1435.5 ± 750.3 | 1518.2 ± 945.6 | 3732.9 ± 2162.6 ###$$$ |
| Morning steps/total steps (%) | 41.2 ± 6.4 | 30.1 ± 5.7 *** | 25.2 ± 6.9 ***& |
| Afternoon steps/total steps (%) | 36.0 ± 5.8 | 46.4 ± 7.0 *** | 30.8 ± 6.8 *&&& |
| Night steps/total steps (%) | 21.0 ± 7.9 | 22.4 ± 7.2 | 42.3 ± 9.4 ***&&& |
| Total MVPA (min/day) | 52.0 ± 24.4 | 47.7 ± 25.2 | 66.0 ± 25.2 & |
| Morning MVPA (min/day) | 23.1 ± 12.2 | 13.9 ± 7.0 *** | 16.0 ± 6.5 * |
| Afternoon MVPA (min/day) | 16.6 ± 8.7 | 20.9 ± 10.1 | 18.2 ± 8.3 |
| Night MVPA (min/day) | 11.1 ± 6.6 | 12.1 ± 9.3 | 30.7 ± 16.1 ###$$$ |
| Morning MVPA/total MVPA (%) | 44.5 ± 5.7 | 30.2 ± 6.7 *** | 25.6 ± 8.3 ***& |
| Afternoon MVPA/total MVPA (%) | 31.2 ± 6.2 | 44.8 ± 7.2 *** | 27.5 ± 6.3 &&& |
| Night MVPA/total MVPA (%) | 22.0 ± 8.9 | 23.5 ± 8.3 | 45.1 ± 8.7 ***&&& |
| **Sleep and meal time** | | | |
| Wake-up time (hh:mm) | 6:14 ± 0:48 | 6:55 ± 0:58 ** | 7:31 ± 1:06 *** |
| Bedtime (hh:mm) | 23:28 ± 0:57 | 0:08 ± 1:09 * | 0:39 ± 0:57 *** |
| Breakfast (hh:mm) | 7:22 ± 0:43 | 7:41 ± 0:45 | 7:54 ± 0:42 |
| Lunch (hh:mm) | 12:29 ± 0:26 | 12:31 ± 0:32 | 12:37 ± 0:27 |
| Dinner (hh:mm) | 19:44 ± 0:49 | 19:55 ± 1:23 | 20:05 ± 0:46 |
| **Dietary intake** | | | |
| Total energy intake (kcal/day) | 2150.3 ± 340.6 | 2260.6 ± 429.3 | 2232.4 ± 331.1 |
| Protein (g/day) | 87.3 ± 20.2 | 88.3 ± 19.1 | 87.2 ± 20.5 |
| Fat (g/day) | 75.0 ± 16.6 | 77.2 ± 20.8 | 78.2 ± 20.4 |
| Carbohydrate (g/day) | 251.6 ± 59.7 | 275.2 ± 67.0 | 256.7 ± 50.7 |
| Dietary fiber (g/day) | 13.9 ± 4.8 | 14.5 ± 4.1 | 12.6 ± 3.8 |

All data are presented as the mean ± standard deviation. BMI: body mass index, MVPA: moderate-to-vigorous physical activity. *** $p < 0.001$, ** $p < 0.01$, * $p < 0.05$ vs morning (One-way ANOVA with Bonferroni), &&& $p < 0.001$, & $p < 0.05$ vs afternoon (One-way ANOVA with Bonferroni), ### $p < 0.001$ vs morning (One-way ANOVA with Dunnett), $$$ $p < 0.001$ vs afternoon (One-way ANOVA with Dunnett).

Supplementary table 6. Physical characteristics of participants by grouping according to the MVPA in males.

| Male (n = 42) | Grouping by MVPA | | |
|---|---|---|---|
| | **Morning (n = 14)** | **Afternoon (n = 19)** | **Night (n = 9)** |
| **Physical characteristics** | | | |
| Age (years) | 46.0 ± 10.6 | 40.2 ± 11.2 | 32.2 ± 10.7 * |
| Height (cm) | 169.9 ± 5.0 | 172.9 ± 5.5 | 173.0 ± 5.5 |
| Weight (kg) | 72.3 ± 12.9 | 71.4 ± 14.0 | 64.8 ± 6.9 |
| BMI (kg/m2) | 25.0 ± 3.6 | 23.8 ± 4.3 | 21.6 ± 1.9 * |
| **Physical activity level** | | | |
| Total step count (steps/day) | 7546.4 ± 4624.0 | 6275.2 ± 2344.4 | 8919.3 ± 4162.9 |
| Morning (steps/day) | 3415.8 ± 2525.8 | 1839.2 ± 625.2 * | 1965.0 ± 728.5 |
| Afternoon (steps/day) | 3014.4 ± 1544.8 | 2707.3 ± 1011.4 | 2569.9 ± 1086.9 |
| Night (steps/day) | 1172.8 ± 613.9 | 1548.7 ± 1070.5 | 3996.3 ± 2513.3 $^{\#\$}$ |
| Morning steps/total steps (%) | 44.0 ± 4.8 | 30.5 ± 7.3 *** | 24.1 ± 7.2 ***$^{\&}$ |
| Afternoon steps/total steps (%) | 36.0 ± 6.6 | 45.5 ± 8.0 ** | 31.8 ± 6.7 $^{\&\&\&}$ |
| Night steps/total steps (%) | 17.6 ± 7.3 | 22.7 ± 8.1 | 42.6 ± 10.1 ***$^{\&\&\&}$ |
| Total MVPA (min/day) | 52.6 ± 31.7 | 45.9 ± 27.5 | 67.4 ± 30.5 |
| Morning MVPA (min/day) | 25.0 ± 17.7 | 13.1 ± 8.3 * | 14.2 ± 6.6 |
| Afternoon MVPA (min/day) | 17.0 ± 11.3 | 20.2 ± 11.3 | 20.4 ± 10.4 |
| Night MVPA (min/day) | 8.6 ± 5.6 | 11.9 ± 9.9 | 31.9 ± 17.6 $^{\#\$}$ |
| Morning MVPA/total MVPA (%) | 46.5 ± 5.2 | 29.6 ± 8.4 *** | 22.7 ± 7.4 *** |
| Afternoon MVPA/total MVPA (%) | 31.7 ± 7.3 | 45.3 ± 9.4 *** | 29.6 ± 6.6 $^{\&\&\&}$ |
| Night MVPA/total MVPA (%) | 18.8 ± 10.1 | 23.7 ± 10.1 | 46.0 ± 7.4 ***$^{\&\&\&}$ |
| **Sleep and meal time** | | | |
| Wake-up time (hh:mm) | 6:10 ± 0:46 | 6:48 ± 1:16 | 7:53 ± 1:34 ** |
| Bedtime (hh:mm) | 23:25 ± 0:56 | 0:16 ± 1:18 * | 0:55 ± 1:04 ** |
| Breakfast (hh:mm) | 7:06 ± 0:35 | 7:31 ± 0:49 | 7:57 ± 0:49 * |
| Lunch (hh:mm) | 12:28 ± 0:25 | 12:24 ± 0:26 | 12:41 ± 0:35 |
| Dinner (hh:mm) | 19:42 ± 0:45 | 20:03 ± 1:16 | 20:04 ± 0:33 |
| **Dietary intake** | | | |
| Total energy intake (kcal/day) | 2526.9 ± 208.6 | 2532.5 ± 442.7 | 2475.4 ± 252.8 |
| Protein (g/day) | 102.4 ± 21.3 | 97.5 ± 20.5 | 98.0 ± 20.8 |
| Fat (g/day) | 84.8 ± 17.2 | 86.8 ± 23.8 | 88.8 ± 20.9 |
| Carbohydrate (g/day) | 301.7 ± 56.6 | 301.2 ± 82.8 | 270.1 ± 66.4 |
| Dietary fiber (g/day) | 16.0 ± 6.2 | 16.3 ± 5.0 | 12.5 ± 3.7 |

All data are presented as the mean ± standard deviation. BMI: body mass index, MVPA: moderate-to-vigorous physical activity. *** $p < 0.001$, ** $p < 0.01$, * $p < 0.05$ vs morning (One-way ANOVA with Bonferroni), $^{\&\&\&}$ $p < 0.001$, $^{\&}$ $p < 0.05$ vs afternoon (One-way ANOVA with Bonferroni), $^{\#}$ $p < 0.05$ vs morning (One-way ANOVA with Dunnett), $^{\$}$ $p < 0.05$ vs afternoon (One-way ANOVA with Dunnett).

Supplementary table 7. Physical characteristics of participants by grouping according to the MVPA in females.

| Female (n = 60) | Grouping by MVPA | | |
|---|---|---|---|
| | **Morning (n = 26)** | **Afternoon (n = 23)** | **Night (n = 11)** |
| **Physical characteristics** | | | |
| Age (years) | 44.9 ± 11.2 | 34.7 ± 8.6 ## | 36.5 ± 15.6 |
| Height (cm) | 160.6 ± 5.0 | 160.7 ± 5.7 | 160.2 ± 4.7 |
| Weight (kg) | 54.2 ± 7.4 | 58.6 ± 11.9 | 57.3 ± 8.7 |
| BMI (kg/m2) | 21.0 ± 2.7 | 22.8 ± 5.0 | 22.4 ± 3.8 |
| **Physical activity level** | | | |
| Total step count (steps/day) | 6835.3 ± 2073.2 | 6447.5 ± 2009.7 | 8015.3 ± 2674.3 |
| Morning (steps/day) | 2710.2 ± 1889.8 | 1889.8 ± 547.5 ** | 2032.0 ± 653.0 * |
| Afternoon (steps/day) | 2466.1 ± 781.6 | 2996.9 ± 803.1 | 2309.8 ± 757.8 |
| Night (steps/day) | 1571.7 ± 788.2 | 1493.0 ± 852.8 | 3517.3 ± 1928.5 ***&&& |
| Morning steps/total steps (%) | 39.7 ± 6.8 | 29.7 ± 4.1 *** | 26.2 ± 6.9 *** |
| Afternoon steps/total steps (%) | 36.1 ± 5.5 | 47.1 ± 6.2 *** | 30.0 ± 7.2 *&&& |
| Night steps/total steps (%) | 22.8 ± 7.8 | 22.2 ± 6.4 | 42.1 ± 9.3 ***&&& |
| Total MVPA (min/day) | 51.7 ± 20.3 | 49.2 ± 23.5 | 64.9 ± 21.4 |
| Morning MVPA (min/day) | 22.1 ± 8.4 | 14.6 ± 5.7 ** | 17.5 ± 6.2 |
| Afternoon MVPA (min/day) | 16.3 ± 7.3 | 21.5 ± 9.3 | 16.4 ± 6.0 |
| Night MVPA (min/day) | 12.4 ± 6.8 | 12.3 ± 9.0 *** | 29.7 ± 15.6 &&& |
| Morning MVPA/total MVPA (%) | 43.4 ± 5.7 | 30.7 ± 4.9 ### | 28.0 ± 8.6 ### |
| Afternoon MVPA/total MVPA (%) | 30.9 ± 5.6 | 44.5 ± 4.9 *** | 25.8 ± 5.8 *&&& |
| Night MVPA/total MVPA (%) | 23.7 ± 8.0 | 23.3 ± 6.8 | 44.4 ± 10.0 ***&&& |
| **Sleep and meal time** | | | |
| Wake-up time (hh:mm) | 6:16 ± 0:49 | 7:00 ± 0:38 ** | 7:13 ± 0:13 ** |
| Bedtime (hh:mm) | 23:30 ± 0:59 | 0:02 ± 1:01 | 0:26 ± 0:50 |
| Breakfast (hh:mm) | 7:33 ± 0:44 | 7:49 ± 0:41 | 7:52 ± 0:37 |
| Lunch (hh:mm) | 12:29 ± 0:28 | 12:37 ± 0:35 | 12:34 ± 0:20 |
| Dinner (hh:mm) | 19:45 ± 0:52 | 19:54 ± 1:03 | 20:05 ± 0:56 |
| **Dietary intake** | | | |
| Total energy intake (kcal/day) | 1955.0 ± 202.0 | 2047.9 ± 275.5 | 2033.5 ± 246.0 |
| Protein (g/day) | 79.5 ± 14.6 | 81.1 ± 14.6 | 78.3 ± 16.2 |
| Fat (g/day) | 70.0 ± 14.1 | 69.6 ± 14.7 | 69.6 ± 16.1 |
| Carbohydrate (g/day) | 225.6 ± 42.7 | 254.9 ± 43.4 | 245.7 ± 32.6 |
| Dietary fiber (g/day) | 12.8 ± 3.6 | 13.0 ± 2.6 | 12.6 ± 4.0 |

All data are presented as the mean ± standard deviation. BMI: body mass index, MVPA: moderate-to-vigorous physical activity. *** $p < 0.001$, ** $p < 0.01$, * $p < 0.05$ vs morning (One-way ANOVA with Bonferroni), &&& $p < 0.001$ vs afternoon (One-way ANOVA with Bonferroni), ### $p < 0.001$, ## $p < 0.01$ vs morning (One-way ANOVA with Dunnett).

Supplementary table 8. Comparison of the glucose parameters among the groups.

| | **Total n = 102** | | | **Male n = 42** | | | **Female n = 60** | | |
|---|---|---|---|---|---|---|---|---|---|
| **Gruping by number of steps** | **Morning (n=29)** | **Afternoon (n=57)** | **Night (n=16)** | **Morning (n=12)** | **Afternoon (n=23)** | **Night (n=7)** | **Morning (n=17)** | **Afternoon (n=34)** | **Night (n=9)** |
| Mean [mg/dl] | 98.5 ± 8.2 | 100.3 ± 10.4 | 94.4 ± 8.2 | 99.4 ± 9.9 | 100.5 ± 7.4 | 99.2 ± 6.7 | 97.8 ± 7.0 | 100.1 ± 12.0 | 90.7 ± 7.4 [&] |
| Standard deviation [mg/dl] | 14.2 ± 4.1 | 14.8 ± 5.3 | 13.9 ± 4.9 | 14.4 ± 3.9 | 14.6 ± 4.9 | 11.2 ± 3.1 | 14.1 ± 4.3 | 14.9 ± 5.7 | 16.0 ± 5.1 |
| CONGA [-] | 5.07 ± 0.45 | 5.17 ± 0.53 | 4.88 ± 0.46 | 5.13 ± 0.57 | 5.18 ± 0.41 | 5.22 ± 0.35 | 5.03 ± 0.35 | 5.16 ± 0.61 | 4.62 ± 0.37 |
| J-INDEX [-] | 12.8 ± 2.1 | 13.4 ± 3.5 | 11.8 ± 2.0 | 13.0 ± 2.5 | 13.3 ± 2.1 | 12.2 ± 2.0 | 12.6 ± 1.9 | 13.5 ± 4.2 | 11.5 ± 2.1 |
| MAGE [mg/dl] | 38.4 ± 14.0 | 38.6 ± 15.8 | 37.6 ± 19.5 | 37.7 ± 12.1 | 39.0 ± 12.4 | 29.0 ± 9.5 | 38.9 ± 15.7 | 38.3 ± 17.8 | 44.3 ± 23.0 |
| MAX [mg/dl] | 138.3 ± 16.7 | 140.3 ± 20.5 | 132.5 ± 18.0 | 137.4 ± 15.7 | 140.8 ± 18.6 | 128.8 ± 13.8 | 139.0 ± 18.0 | 140.0 ± 21.9 | 135.4 ± 21.0 |
| MIN [mg/dl] | 79.3 ± 8.7 | 79.9 ± 10.2 | 75.7 ± 10.4 | 79.2 ± 11.4 | 81.3 ± 9.9 | 82.9 ± 5.7 | 79.4 ± 6.3 | 79.0 ± 10.4 | 70.2 ± 9.8 [&] |
| **Grouping by MVPA** | **Morning (n=40)** | **Afternoon (n=42)** | **Night (n=20)** | **Morning (n=14)** | **Afternoon (n=19)** | **Night (n=9)** | **Morning (n=26)** | **Afternoon (n=23)** | **Night (n=11)** |
| Mean [mg/dl] | 100.1 ± 10.8 | 98.6 ± 8.9 | 96.8 ± 8.7 | 98.8 ± 9.5 | 100.5 ± 7.4 | 100.5 ± 6.9 | 100.8 ± 11.6 | 97.0 ± 9.8 | 93.7 ± 9.2 |
| Standard deviation [mg/dl] | 16.1 ± 5.7 | 13.4 ± 4.0 | 13.8 ± 4.4 | 15.3 ± 4.7 | 13.6 ± 4.4 | 12.7 ± 4.2 | 16.5 ± 6.2 | 13.2 ± 3.7 | 14.7 ± 4.6 |
| CONGA [-] | 5.14 ± 0.54 | 5.10 ± 0.50 | 4.99 ± 0.48 | 5.10 ± 0.55 | 5.19 ± 0.41 | 5.26 ± 0.33 | 5.17 ± 0.54 | 5.03 ± 0.55 | 4.78 ± 0.49 |
| J-INDEX [-] | 13.7 ± 3.8 | 12.6 ± 2.3 | 12.3 ± 2.1 | 13.1 ± 2.4 | 13.1 ± 2.1 | 12.9 ± 2.2 | 14.0 ± 4.4 | 12.2 ± 2.4 | 11.8 ± 2.1 |
| MAGE [mg/dl] | 42.5 ± 17.3 | 34.9 ± 12.4 | 37.4 ± 17.7 | 38.1 ± 12.5 | 38.3 ± 12.7 | 32.4 ± 10.7 | 44.9 ± 19.2 | 32.1 ± 11.7 | 41.4 ± 21.6 |
| MAX [mg/dl] | 143.9 ± 20.8 | 135.4 ± 17.4 | 134.8 ± 17.6 | 140.3 ± 19.2 | 137.6 ± 16.8 | 134.5 ± 16.5 | 145.8 ± 21.8 | 133.5 ± 18.1 | 135.0 ± 19.2 |
| MIN [mg/dl] | 78.8 ± 9.0 | 79.7 ± 10.7 | 78.5 ± 9.8 | 78.2 ± 11.9 | 81.8 ± 9.5 | 83.5 ± 5.2 | 79.1 ± 7.2 | 77.9 ± 11.5 | 74.3 ± 10.9 |

All data are presented as the mean ± standard deviation. CONGA: continuous overall net glycemic action, MAGE: mean amplitude of glycaemic excursion, MAX: maximum glucose, MIN: minimum glucose. [&] $p < 0.05$ vs afternoon (One-way ANOVA with Bonferroni).
